# Detecting HIV positive cases among first-time testers using HIV self-testing strategy: evidence from a large-scale cross-sectional study in Cameroon

**DOI:** 10.1101/2025.05.20.25328024

**Authors:** Bouba Yagai, Jeremiah Efakika Gabisa, Audrey Raissa Djomo Dzaddi, Fatima Mouliom Nkain, Adamou Souleymanou, Albert Franck Zeh Meka, Yacouba Liman, Annie Michele Salla, Lily Claire Ekobika, Gutenberg Tchikangni, Alex Durand Nka, Aude Christelle Ka’e, Antoine Socpa, Rogers Awoh Ajeh, Marie José Essi, Hadja Cherif Hamsatou, Anne Cecile Zoung-Kanyi Bissek, Joseph Fokam, Serge Clotaire Billong

**Affiliations:** Chantal BIYA International Reference Centre for research on HIV/AIDS prevention and management (CIRCB), Yaounde, Cameroon; UniCamillus - Saint Camillus International University of Health Sciences, Rome, Italy; Health Office, United States Agency for International Development (USAID), Yaounde, Cameroon; Division of Health Operational Research, Ministry of Public Health, Yaounde, Cameroon; University of Yaounde I, Yaounde; Central Technical Group, National AIDS Control Committee, Yaounde, Cameroon; Association Camerounaise pour le Marketing Social (ACMS), Yaounde, Cameroon; University of Buea, Buea, Cameroon

**Keywords:** HIV, first-time testers, HIV self-testing, seropositivity, distribution models, Cameroon

## Abstract

**Introduction:** To end AIDS by 2030, barriers in accessing routine HIV testing services must be addressed, especially in high-risk populations. Innovative strategies such as HIV self-testing (HIVST) represent an opportunity to overcome the existing gaps. We aimed at characterizing HIV first-time testers (HFTs) and evaluating the effectiveness of HIVST among HFTs.

**Methods:** A cross-sectional study was conducted in three regions of Cameroon. Oral HIVST kits were distributed using four distribution models: community model (CM), workplace model (WM), partners of PLHIV model (pPLHIVM), and other health facility entry points model (oHFM).

**Results:** Overall, 6018 HFTs were analysed; 69.8% were males (male/female ratio: 2.3) and 82.2% were <30 years (<20 years: 26.4%, 20-29 years: 55.7%). Most kits were distributed using primary distribution type (78.1%) and CM (69.7%). Concerning HIVST outcomes, 2.3% (n=137) and 0.7% (n=39) had reactive and invalid test results, respectively. Of them, 45.5% did not show up for confirmation; most of them had <20 years (78.0%) and received their kits through primary distribution (73.6%). However, a concordance of 77.1% was found between the results of HIVST and health-care workers (HCW); the lowest rate was observed in <20 years (55.6%). The overall HIV seropositivity rate was 1.2%. Compared to individuals aged 40-49 years (4.6%), those <20 years and 20-29 years had a seroprevalence <1%, p<0.001. pPLHIVM showed the highest prevalence (18.2%), compared to CM (0.1%), p<0.001. Using the conditional backward stepwise regression model, predictors of seropositivity were region, distribution type, and distribution models. Compared to primary distribution, secondary distribution had 51 times odd of positive test (aOR [95% CI]: 51.447 [4.627-572.010]).

**Conclusions:** About 70% of HFTs were males and 80% had <30 years. Only 54% showed up for confirmation testing, but a good concordance (77%) of test results between HFTs and HCWs was observed. HIV seropositivity was low, with a much higher rate among partners of PLHIV. This real-life data suggests that HIVST could play an important role in achieving the first 95% UNAIDS targets, especially among males and young people.

## Introduction

Several decades after its discovery, HIV/AIDS remains a significant public health threat, but efforts have been made to curb HIV transmission. Factors contributing to the reduction in transmission and global spread include increased awareness and education, access to antiretroviral therapy, harm reduction strategies, safe sex promotion, improved testing and counseling services, and targeted interventions for key populations [1,2]. The global number of HIV new infections has reduced significantly by 38%, from 2.1 million [1.6 million–2.8 million] in 2010 [3] to 1.3 million [1.0 million–1.7 million] in 2022 [4]. However, the UNAIDS 2022 report highlights challenges in achieving the 2030 AIDS target; with approximately 4000 new HIV cases daily, including 1100 individuals aged of 15- 24 [5]. If the present trends persist, there could be 1.2 million new cases of HIV in 2025 [5]. In Cameroon, progress towards the 95-95-95 target is notable but some challenges still remain in several areas. The National AIDS control committee 2021 annual report indicates that only 83.2% of PLHIV know their status, 84.9% have access to ART, and 84.5% are virally suppressed [6]. Therefore, addressing gaps in testing and treatment is crucial to achieving the 95% UNAIDS targets by 2025.

Cameroon still faces some challenges in its HIV testing system, despite advancements in HIV testing technologies [7], including limited access to existing HIV testing services, stigma and discrimination, and limited awareness on HIV testing in some populations [2,8]. In the path to ending AIDS by 2030, global communities should consider new and innovative strategies that can help overcome these testing barriers. One of these approaches recommended by WHO since 2016 is the HIV self-testing (HIVST) strategy, which can reach key populations, men, and young people who may not test otherwise [9]. Many sub-Saharan countries have adopted the HIVST, but the result of its implementation remains generally poorly documented.

HIVST is a process whereby people collect their own sample, perform their HIV rapid diagnostic test in a private setting, either alone or with a trusted person, and interprets the result. Studies have shown that HIVST can be comparable or is even more cost-effective compared to standard HIV testing methods [10–12], especially when implemented by the community. It is also interesting to note that self-testing strategy is generally accepted by most potential target populations [13,14] [14–18] and policy makers [19]. Such new approach could therefore play an important role in increasing testing and ART uptake in Africa [20–22], with the final aim of achieving the UNAIDS 95-95-95 targets by 2025.

Data from the Self Testing Africa Initiative (STAR) in Cameroon showed that about 16.7% (6032/35934/) of participants who received the kits were first-time testers [23]. In this study, we aimed to address the following important questions: (i) what is the socio-demographic characteristic of the HIV first-time testers reached by HIVST? (ii) How effective is HIVST in detecting new HIV cases among first-time testers and linking them to ART? The results of this study might provide useful information to public health programs and policy makers in designing an appropriate strategy to reach first-time HIV testers.

## Methods

### Study design and populations

A descriptive cross-sectional study with an analytical design was carried out in the Littoral (economic capital), Centre (political capital), and the South (with the highest HIV prevalence of 5.8%, according to DHS 2018 report [8]) regions of Cameroon, representing the heterogeneity of the country.

### Distribution models

HIV test kits were distributed either directly to the people concerned (primary distribution) or through a third party (secondary distribution). Three main distribution models were used to deliver HIV self-testing kits: community, workplace, and health facility-based models. The health facility-based model was further divided into partners of PLHIV model (excluding HIV+ pregnant women), and other entry points. or at the HIV testing sites.

### Test kits, sample types and testing algorithm

The test kit used in the HIVST was Oraquick HIV-1/2 (OraSure Technologies Inc., Bethlehem, Pennsylvania, United States), which an oral self-testing method. The test kit containing the step-by-step instructions on the procedure to perform and interpret the test, and test device was given to each participant (https://uk.oraquick.com/media/wysiwyg/OQHIVST_CE_IFU_-_English.pdf). In case of a reactive or invalid result, a confirmatory test was performed in a health facility following the national HIV testing algorithm. The confirmatory testing was carried out according to the national guideline, which was based on two tests serial algorithm of rapid test, with blood as samples (whole blood, plasma or serum).

The tests were interpreted according to the manufacturer’s instructions and as per the national procedures as follows: (i) “invalid” when the “control” band was absent, regardless of the presence of a band in the “test” area. (ii) “non-reactive” when the “control” band was present but absent in the “test” area. (iii) “reactive” when both the “control” and “test” bands were present, regardless of band intensity. With regards to the final participants’ serological status, the results were interpreted as follows: (i) “Negative” when the first test is “non-reactive”. In this case, there is no need to proceed to the second test. (ii) “Positive” where both the first and second tests were “reactive”. (iii) “Inconclusive” where the first test was “reactive” but the second test was “non-reactive”. In this case, a tie breaker test was performed according to the national HIV testing algorithm.

### Data collection and analysis

Trained personnel in companies, health care workers, and community mobilization agents were responsible for distributing the kits and collecting the data, which were then entered into the national database (DHIS2) after a data validation process. After data compilation, it was extracted from DHIS2 and cleaned; then exported to SPSS version 26 (SPSS Inc., Chicago, IL, USA). In the present study, only the data of participants who self-reported to have never performed an HIV test before (routine or self-test) was considered. Data were summarized as proportions using tables and figures. The proportions among the various categories of the variables of interest were compared using the Chi-square test or Fisher’s Exact test as appropriate. Binary regression model was used to identify factors independently associated with presenting for confirmation test, concordance in test results and HIV positivity among first-time testers. All the variables in the univariate analysis were fitted and adjusted in the conditional backward stepwise regression model. The threshold for statistical significance was set at p<0.05 at the univariate level. The Hosmer and Lemeshow test and Nagelkerke R^2^ were used for the evaluation of the goodness-of-fit.

### Ethical considerations

This study was conducted in accordance with the principles of the Declaration of Helsinki and national regulations.

Participation was voluntary and all participants enrolled either through the primary kit distribution type granted verbal consent for participation directly, while a third-party verbal consent for those involved through the secondary kit distribution type was provided by the third party involved.

The study protocol was approved by the Institutional Ethical Review Board of the Faculty of Medicine and Biomedical Sciences (FMBS) of the University of Yaoundé 1 (Reference No. 0024/UY1/FMSB/VDRC/CSD) on September 13, 2021. Moreover, all the required administrative authorizations at both the national and regional levels were obtained. All the information was entered into the DHIS2 system, which is a national information system in which data are codified, with restricted access to persons unauthorised.

## Results

### Characteristics of study participants

Of 6019, a total of 6018 first-time HIV testers were considered for the present analysis (one excluded due to incomplete data), with most of them (N=4539) enrolled in the Centre region (Table 1). Most of the participants were males (69.8%), with a male to female ratio of 2.3. Concerning the age structure, the majority were in the age group 20-29 years (55.7%), followed by <20 years (26.4%). Regarding the proportions of males and females according to age groups, females were significantly higher in the group <20 years (39.9%), when compared to males (20.6%, p<0.001). While no difference between the proportions of males and females in the 20- 29 years group, as from 30 years, females were significantly lower when compared to males (Figure 1). Most participants (78.1%) received their test kits through primary distribution type. Concerning the distribution model employed, most kits were distributed through the community model (69.7%), followed by other HF entry point model (17.8%).

**Table 1.**
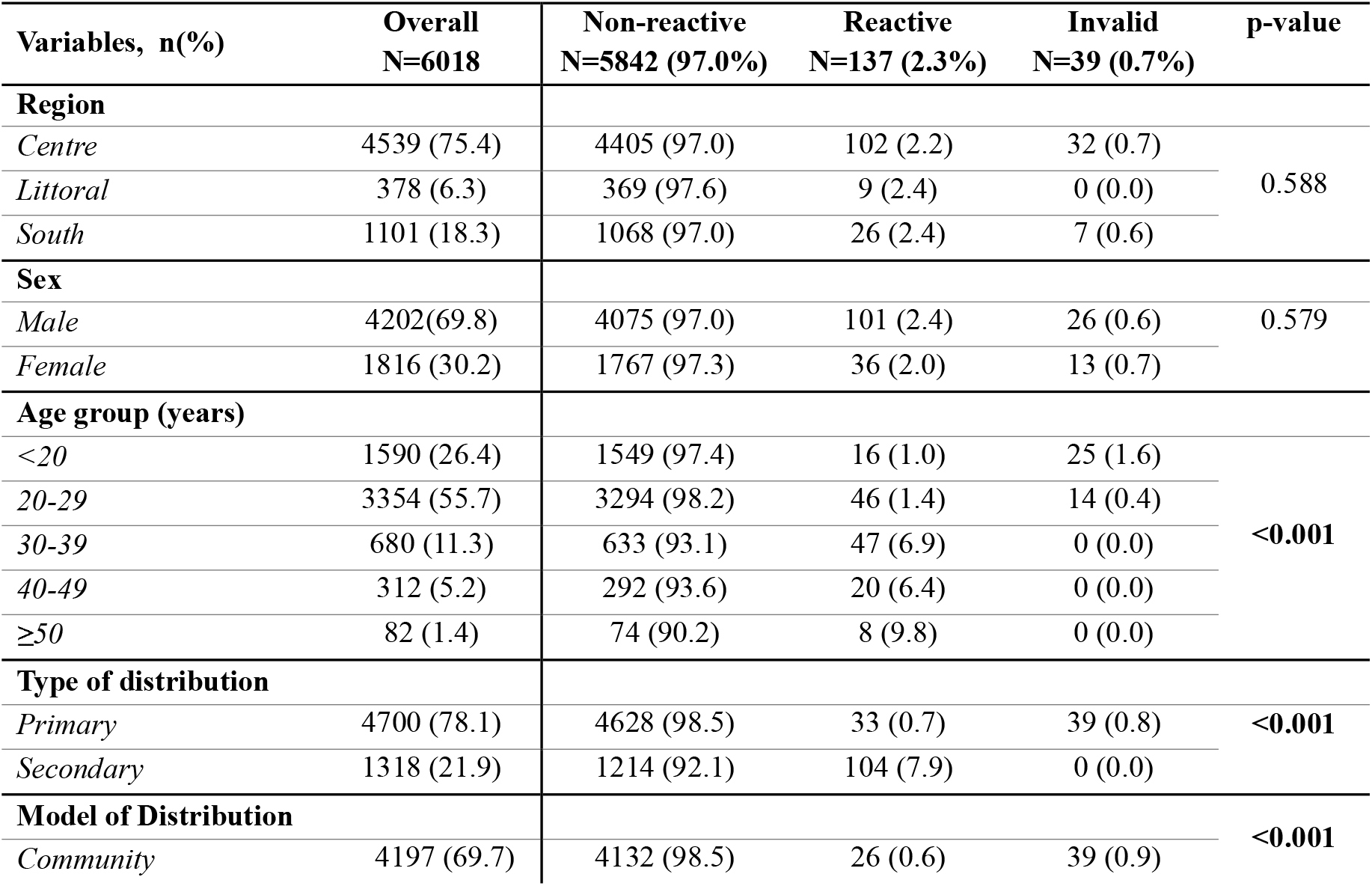
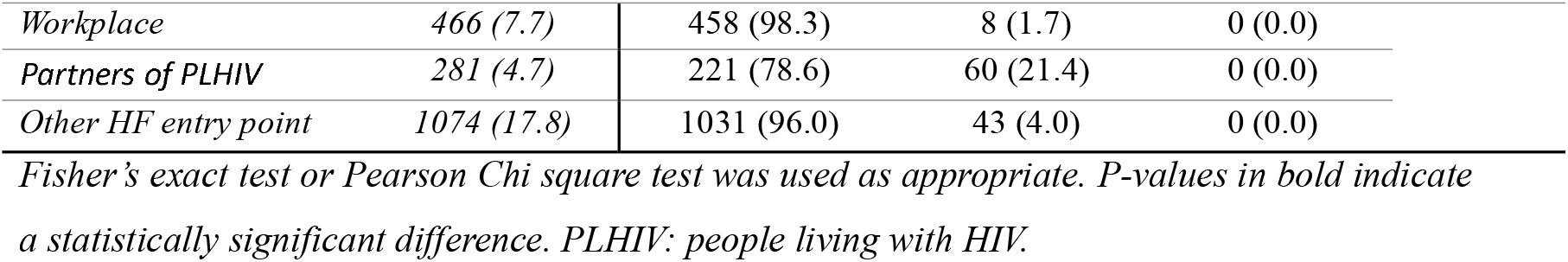
HIV self-testing outcome among first-time testers.

| Variables, n(%) | Overall<br>N=6018 | Non-reactive<br>N=5842 (97.0%) | Reactive<br>N=137 (2.3%) | Invalid<br>N=39 (0.7%) | p-value |
| --- | --- | --- | --- | --- | --- |
| **Region** |  |  |  |  |  |
| *Centre* | 4539 (75.4) | 4405 (97.0) | 102 (2.2) | 32 (0.7) | 0.588 |
| *Littoral* | 378 (6.3) | 369 (97.6) | 9 (2.4) | 0 (0.0) |  |
| *South* | 1101 (18.3) | 1068 (97.0) | 26 (2.4) | 7 (0.6) |  |
| **Sex** |  |  |  |  |  |
| *Male* | 4202(69.8) | 4075 (97.0) | 101 (2.4) | 26 (0.6) | 0.579 |
| *Female* | 1816 (30.2) | 1767 (97.3) | 36 (2.0) | 13 (0.7) |  |
| **Age group (years)** |  |  |  |  |  |
| <20 | 1590 (26.4) | 1549 (97.4) | 16 (1.0) | 25 (1.6) | <0.001 |
| 20-29 | 3354 (55.7) | 3294 (98.2) | 46 (1.4) | 14 (0.4) |  |
| 30-39 | 680 (11.3) | 633 (93.1) | 47 (6.9) | 0 (0.0) |  |
| 40-49 | 312 (5.2) | 292 (93.6) | 20 (6.4) | 0 (0.0) |  |
| $\geq 50$ | 82 (1.4) | 74 (90.2) | 8 (9.8) | 0 (0.0) | |
| **Type of distribution** |  |  |  |  |  |
| *Primary* | 4700 (78.1) | 4628 (98.5) | 33 (0.7) | 39 (0.8) | <0.001 |
| *Secondary* | 1318 (21.9) | 1214 (92.1) | 104 (7.9) | 0 (0.0) |  |
| **Model of Distribution** |  |  |  |  |  |
| *Community* | 4197 (69.7) | 4132 (98.5) | 26 (0.6) | 39 (0.9) | <0.001 |

|  |  |  |  |  |
| --- | --- | --- | --- | --- |
| *Workplace* | 466 (7.7) | 458 (98.3) | 8 (1.7) | 0 (0.0) |
| *Partners of PLHIV* | 281 (4.7) | 221 (78.6) | 60 (21.4) | 0 (0.0) |
| *Other HF entry point* | 1074 (17.8) | 1031 (96.0) | 43 (4.0) | 0 (0.0) |

*Fisher's exact test or Pearson Chi square test was used as appropriate. P-values in bold indicate* *a statistically significant difference. PLHIV: people living with HIV.*

**Figure 1.**
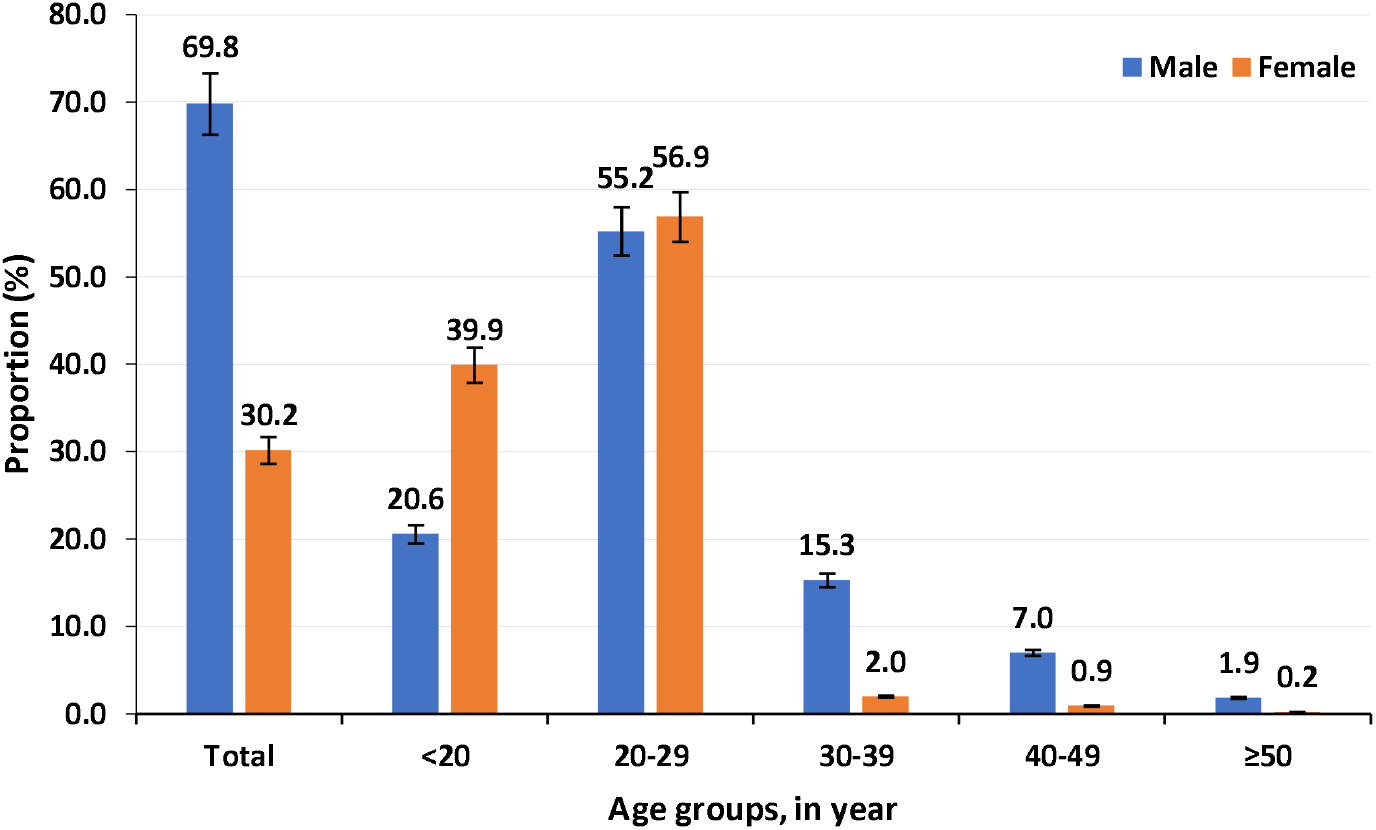
Proportions of male and female first-time testers according to age groups.

### HIV self-testing outcome among first-time testers

The outcome of self-testing is reported in Table 1. Globally, a total of 137 individuals had a reactive test (2.3%); while 0.7% had invalid test results. The proportions of non-reactive and reactive results were closely similar across the three regions and across sex (Table 1).

According to age groups, those ≥50 years had the highest proportion of reactive tests (9.8%), followed by 30-39 years (6.9%), 40-49 years (6.4%), 20-29 years (1.4%), and finally <20 years (1.0%), p<0.001. A significantly higher proportion of reactive tests was observed in the secondary distribution type, when compared to primary distribution (7.9% versus 0.7%), p<0.001. Looking at the distribution models, the partners of PLHIV model showed the highest proportion of reactive tests (21.4%), followed by other HF entry point model (4.0%); while the community model showed only 0.6% reactive tests, p<0.001. Interestingly, the proportion of invalid tests was small (<1%), and all those with invalid test results were those aged <30 years (Table 1).

Of note, only 54.5% (96/176) of individuals with results requiring confirmation at health facility level showed up, the majority of those aged <20 years (78.0%), and those ≥50 years (62.5%) did not show up for confirmation. Primary distribution type and the PLHIV model was a risk factor and an enabling factor for confirmatory testing, respectively. Similarly, among those who showed up for confirmation, 77.1% of results were concordant with those of health facility-based findings. A high discordance rate was observed in the primary distribution type (57.9%) and community distribution model (64.3%); while a high concordance was observed in the partners of PLHIV model (94.3%).

### HIV seropositivity and ART Linkage among first-time self-testers

The overall seroprevalence (as confirmed by the national algorithm) was 1.2% (74/5938). A similar prevalence was found in males and females HIV first-time testers (Table 4). Compared to older individuals (40-49 years: 4.6%, 30-39 years: 3.9%, and ≥50 years: 3.9%), younger age <20 and 20-29 years had a significantly lower seroprevalence (0.3% and 0.8%, respectively), p<0.001 (Table 4). The secondary distribution type showed a significantly higher prevalence (5.1%), when compared to primary distribution (0.2%), p<0.001 (Table 4). Regarding the distribution model, the highest prevalence was observed in the partners of PLHIV model (18.2%), followed by other HF entry point model (1.4%). Among the participants in the workplace and community models, the HIV seroprevalence was <1% each.

**Table 4.**
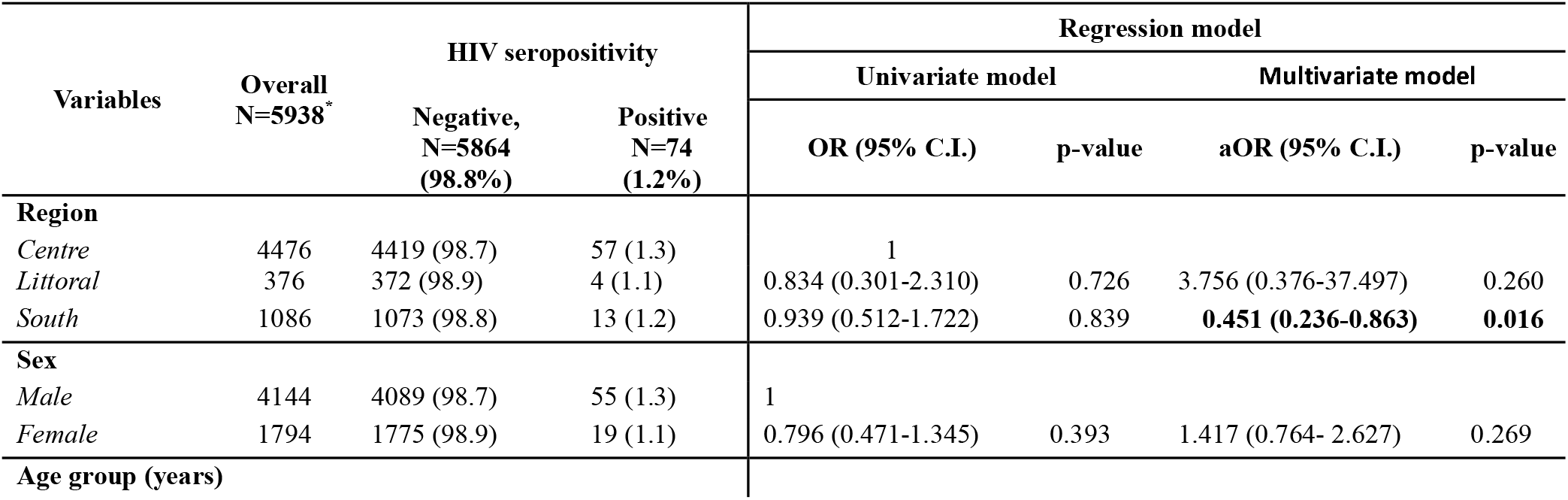
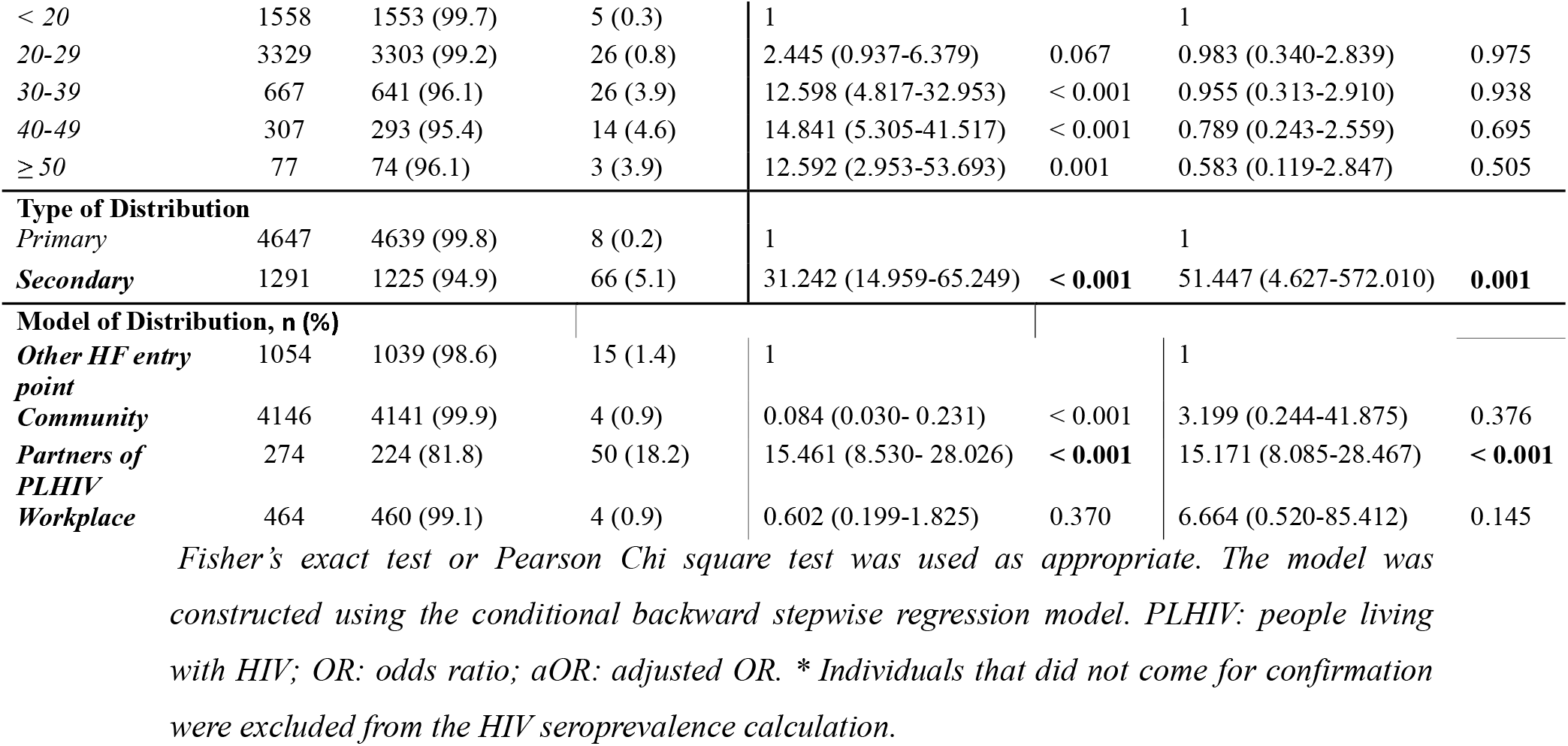
HIV seropositivity and ART linkage among self-testers.

In the regression analysis at the univariate level, age group, distribution type, and distribution model were associated with HIV seropositivity. After adjusting for all variables in the conditional backward stepwise regression model for the goodness-of-fit, the Nagelkerke R^2^ and the Hosmer and Lemeshow tests were 0.358 and < 0.001, in this order. Region, distribution type, and distribution model were independent predictors of HIV seropositivity among first-time testers. In particular, the South region showed about 2 times lesser odds for a positive test, compared to the Centre region (aOR [95% CI]: 0.451 [0.236-0.863]). Compared to the primary distribution type, the secondary distribution type showed about 51 times the odd of positive HIV test (aOR [95% CI]: 51.447 [4.627-572.010], p=0.001). Also, compared to other HF entry point model, the PLHIV model showed about 15 times the odds of HIV positive test (aOR [95% CI]: 15.171 [8.085-28.467]). (Table 4).

Of the 74 confirmed positive cases, all those with available data (n=69) were successfully initiated on ART.

## Discussion

Over time, eliminating barriers to HIV services and ensuring equitable service delivery has been a great concern. The STAR initiative was launched as a response to this issue. Our study evaluated the demographic traits of first-time testers and the effectiveness of HIVST to identify infected individuals among first-time testers and to link them to ART.

Our findings reveal a notable number of individuals in Cameroon who have never undergone HIV testing, indicating persistent barriers for access to testing. Some of the barriers outlined in similar settings include stigma and discrimination, lack of awareness, cultural and language barriers, accessibility, cost, fear, and denial [24–27]. Introducing strategies like self-testing can be important to reach undeserved communities and individuals beyond traditional services. Our analysis shows that, the majority (69.8%) of the first-time HIV self-testers were males, aligning with the 2018 Demographic Health Survey data in Cameroon that indicate a higher proportion of untested men (43%) compared to women (29%) [8]. Existing data indicate that males are less likely to seek care, when compared to females [28–31]. The advent of HIVST offers a unique opportunity for men and women to test for HIV in the privacy of their own homes. Some reports indicates that, HIVST has helped men in Eswatini as well as those in Malawi who had issues with going to health facilities to access testing [32]. It was interesting to note that, even though the majority of study participants in this study were males, females were more frequent in <20 years old, not in subsequent older age group. This indicates the need to design strategies to reach young girls with testing. This could be done by integration of services designed for them to reduce the existing gaps. Over 80% of our study participants were below 30 years. The community and workplace distribution models have been demonstrated to primarily target young people, underlining the suitability of self-testing for this demographic, shown to include key populations like sex workers and injection drug users [33,34]. These results align with the observation that, young adults (up to 75%) prefer HIVST [32].

In our analysis, as an outcome of self-testing among first-time testers, only 2.3% and <1% showed an invalid result. As indicated in other settings, this low proportion of invalid results might indicate the fact that first-time testers find the self-testing feasible [17,35–37]. Overall, the proportion of those who were eligible for confirmation and showed up was only 54.5%. The observation that none of the 39 first-time testers with invalid test results showed up for confirmation is worrisome and should be appropriately addressed at the counseling step. However, about 70% of first-time testers with reactive tests were confirmed according to the national algorithm. This proportion was especially very low among youths aged <20 years (only about 22%). When designing self-testing strategy, particular attention should be given to youths, especially those who are at higher risk of HIV infection.

Beyond showing up for the confirmation test, our analysis showed that the concordance between self-testers and HCW was good, notably in the model of PLHIV. The overall seroprevalence among HIV first-time testers was lower than the national prevalence which is about 2.7% [38]. This low prevalence might be partly explained by the potentially low HIV risk among the targeted populations in the workplace and community-based models. To be cost effective, it is important to appropriately select the high-risk groups in the communities. As expected, the highest prevalence was observed among partners of PLHIV model. This underscores the importance of achieving and maintaining virological suppression, especially in the case of discordant couples to reduce the risk of HIV transmission [39]. This suggests that the distribution of HIVST kits to the partners of those with unsuppressed viral load could be a potentially effective strategy in resource limited settings. In fact, it was reassuring to observe that first-time testers are best reached by the secondary distribution model. It should be noted that higher prevalence was observed in the PLHIV model in other settings [40]. The extremely low prevalence observed in other models is indicative of the need to adjust the target population for HIVST in the national HIV testing strategy. Even though only a few cases were identified, it was interesting to observe that those who had available follow up data were linked to ART. In addition to contributing to the achievement of the first 95%, our data also suggest a potential role of HIVST strategy in achieving the second 95% among first-time testers [32,41–43]. Despite the good sample size, this study has some limitations. First, the data analyzed here is routine data, not initially collected for research purposes; therefore, it is not free of selection bias. Secondly, due to the few available socio-demographic variables, first-time testers could not be extensively characterized. This perhaps, justifies why the Hosmer and Lemeshow test for the goodness of fit did not fit the seropositivity covariates, demonstrating redundancy in possible multicollinearity. However, to the best of our knowledge, we present for the first-time real-life data on the use of HIVST among HIV first-time testers in Cameroon. Additional studies are warranted to provide an exhaustive description of first-time testers and to confirm the results of this study.

## Conclusion

About 70% and 80% of HIV first-time testers were males and young people <30 years. Even though a good concordance of HIV result between HIVST and HCWs was observed (77%), only about half (54%) showed up for confirmation testing at health facility. Overall, HIV seropositivity among first-time testers was low (1.2%), with a much higher rate among partners of PLHIV. This real-life data suggests that HIVST could play an important role in achieving the first 95% UNAIDS targets, especially among males and young people.

## Author’s contribution

Conceptualization and methodology: BY, SCB, ARDZ, AMS, LCE, GT, AS, RAA, MJE, HHC, AZKB, AFZM, JF, MNF, YL and JEG. Funding acquisition: SCB, AMS, LCE, GT. Data curation, formal analysis: BY, JEG, DDAR, ADN and ACK. Visualization: BY, JEG, ARDD, ADN and ACK. Original draft preparation: BY, JEG, and ARDZ. Review and editing: SCB, JF, AS, RAA, AS, MJE and AFZM. Supervision and project administration: HHC, SCB, AMS, LCE and GT. All the authors read and approved the final version of the manuscript.

## Acknowledgments

We thank ACMS for collaborating to secure funding and for providing technical assistance for the implementation of the project.

## Data Availability Statement

Data is available upon reasonable request from the corresponding author.

